# Evaluation on the diagnostic efficiency of different methods in detecting COVID-19

**DOI:** 10.1101/2020.06.25.20139931

**Authors:** Haitao Yang, Yuzhu Lan, Xiujuan Yao, Sheng Lin, Baosong Xie

## Abstract

**Objective:** To evaluate the diagnostic efficiency of different methods in detecting COVID-19 to provide preliminary evidence on choosing favourable method for COVID-19 detection.

**Methods:** PubMed, Web of Science and Embase databases were searched for identifing eligible articles. All data were calculated utilizing Meta Disc 1.4, Revman 5.3.2 and Stata 12. The diagnostic efficiency was assessed via these indicators including summary sensitivity and specificity, positive likelihood ratio (PLR), negative LR (NLR), diagnostic odds ratio (DOR), summary receiver operating characteristic curve (sROC) and calculate the AUC.

**Results:** 18 articles (3648 cases) were included. The results showed no significant threshold exist. EPlex: pooled sensitivity was 0.94; specificity was 1.0; PLR was 90.91; NLR was 0.07; DOR was 1409.49; AUC=0.9979, Q*=0.9840. Panther Fusion: pooled sensitivity was 0.99; specificity was 0.98; PLR was 42.46; NLR was 0.02; DOR was 2300.38; AUC=0.9970, Q*=0.9799. Simplexa: pooled sensitivity was 1.0; specificity was 0.97; PLR was 26.67; NLR was 0.01; DOR was 3100.93; AUC=0.9970, Q*=0.9800. Cobas^®:^ pooled sensitivity was 0.99; specificity was 0.96; PLR was 37.82; NLR was 0.02; DOR was 3754.05; AUC=0.9973, Q*=0.9810. RT-LAMP: pooled sensitivity was 0.98; specificity was 0.99; PLR was 36.22; NLR was 0.04; DOR was 751.24; AUC=0.9905, Q*=0.9596. Xpert Xpress: pooled sensitivity was 0.99; specificity was 0.97; PLR was 27.44; NLR was 0.01; DOR was 3488.15; AUC=0.9977, Q*=0.9829.

**Conclusions:** These methods (ePlex, Panther Fusion, Simplexa, Cobas^®^, RT-LAMP and Xpert Xpress) bear higher sensitivity and specificity, and might be efficient methods complement to the gold standard.

## Introduction

Severe Acute Respiratory Syndrome Coronavirus 2 (SARS-CoV-2) belonging to the genus beta coronavirus, is the seventh and most novel human coronavirus. It is highly homologous to the sequences of coronaviruses found in humans, bats and other wild animals (like SARS-CoV and bat SARS-like CoV), with a sequence homology of 80% to 89%(1-8)

Corona Virus Disease 2019(COVID-19) often presents with fever, myalgia, cough, dyspnea and atypical imaging findings (9-11). However, there are also cases with asymptomatic or mildly symptomatic(12-15). By 9 June 2020, 7 039 918 confirmed cases have been reported to the World Health Organization (WHO) from 216 countries, with 404 396 deaths(16). COVID-19 has caused a worldwide epidemic and most countries have implemented a containment strategy as advised by the WHO to control further transmission(17, 18). It is urgent to develop rapid, accurate diagnosis methods to effectively identify these early infected patients, treat them in time and control the disease spreading(19).

Through metagenomic sequencing, SARS-CoV-2 was first identified in BALF (bronchoalveolar lavage fluid) of Chinese patients(10, 20, 21). Based on the rapid characterization of the full genome of the virus, many molecular diagnostic methods have been developed rapidly. Real-time reverse transcription-polymerase chain reaction (rRT-PCR) is currently the standard of the diagnosis of acute coronavirus disease (COVID-19)(10, 21, 22). However, rRT-PCR assay has many limitations, such as required high purity samples, trained personnel and time-consuming. This test did not meet the rapidly growing need for virus testing in patients with COVID-19 infection, suspected infection or close contact with confirmed cases, which have significantly hampered public health efforts to contain the outbreak. To resolve this contradiction, the U.S. Food and Drug Administration (FDA) issued Emergency Use Authorization (EUA) for multiple SARS-CoV-2 rapid tests beginning in March 2020. Up to now, some studies have been published to comparing the diagnostic features of these methods(23-25), whereas the results were inconsistent. Therefore, it is urgent to performed this meta-analysis to comprehensively summarize the characteristics of the diagnostic assay in detecting COVID-19 to provide the preliminary evidence to support further guide clinical routine through evidence-based medicine.

## Materials and Methods

### Search strategy

Studies on evaluation of the method for COVID-19 detection in PubMed, Web of Science and Embase databases were thoroughly searched, from their inception till 25 March 2020. The Searching terms include: “PCR”, “COVID-19”, “SARS-CoV-2”, “Corona Virus Disease 2019”, “RT-PCR”, “diagnosis”. The language was limited in English.

### Study selection

Pertinent articles were identified by two reviewers after screening titles or abstracts. Duplicated articles were removed by checking duplication. Besides, references of selected articles were screened to find more relevant articles. Disagreement was resolved by the third reviewer. All eligible articles were evaluated by Quadas-2 for systematic reviews of intervention (Version 5.3.0).

### Inclusion and exclusion criteria

Articles meeting the following criteria were included in this study: i) the articles studied on the method for detecting COVID-19; ii) patients recruited in articles should include COVID-19 and non-COVID-19; iii) patients defined as COVID-19 or non-COVID-19 were proved by gold standard; iv) data in articles were sufficient to analyze the pooled sensitivity and specificity; v) articles published in English. Incomplete data, reviews and case reports were excluded.

### Data extraction

Two reviewers independently pooled the following data from eligible articles: author, year, Country, sample size, methods, true positive (TP), false positive (FP), false negative (FN), true negative (TN). When the extracted data were inconsistent, discrepancies were resolved by discussion or the third reviewer.

### Statistical method

All statistical analyses were conducted by employing Meta-Disc 1.4, RevMan 5.3.2, Stata 12 software. The threshold effect was evaluated by computation of Spearman correlation coefficient between the logit of sensitivity and logit of 1-specificity. A strong positive correlation suggested the threshold effect. Heterogeneity among included articles was assessed via Chi-square and I-square tests. I^2^>50% or *P*<0.1 were deemed as significant heterogeneity existing. The random-effects or fixed-effects model was chosen to analyze the pooled data depending on the presented heterogeneity. The summary sensitivity and specificity, positive likelihood ratio (PLR), negative LR (NLR), diagnostic odds ratio (DOR), summary receiver operating characteristic curve (sROC) and calculate the AUC were utilized to evaluate the diagnostic efficiency of the aboved methods in detecting COVID-19. The publication bias was evaluated by Deek’s funnel plot asymmetry test. *P*>0.1 suggested no significant publication bias in this study.

## Results

After searching three databases, out of 1094 duplicate articles, 3874 articles were identified. Then, 3837 articles were removed by screening the title or abstract. Afterwards, 19 articles were further excluded via thoroughly reading the full-text. Eventually, 18 articles of 3648 cases were included in this meta-analysis. Table 1 described the characteristics of included articles. Supplementary Figure 1 exhibited the progression of searching. Considering various of methods among included articles, we conducted this meta-analysis based on the different methods (3 articles for ePlex(26-28); 4 articles for Panther Fusion(26, 27, 29, 30); 3 articles for Simplexa(26, 29, 31); 3 articles for Cobas^®^(29, 32-34) ; 5 articles for Xpert Xpress(27, 29, 35-37), 6 articles for RT-LAMP(38-43)).

**Table 1.** the characteristics of included studies.

| Author | Nation | sample size | method | TP | FP | FN | TN |
| --- | --- | --- | --- | --- | --- | --- | --- |
| Zhen et al(a), 2020[26] | The USA | 104 | ePlex® | 49 | 0 | 2 | 53 |
| Zhen et al(b), 2020[27] | The USA | 108 | ePlex® | 53 | 0 | 5 | 50 |
| Uhteg et al, 2020[28] | The USA | 68 | ePlex® | 13 | 0 | 0 | 34 |
| Zhen et al(a), 2020[26] | The USA | 104 | Panther Fusion | 51 | 2 | 0 | 51 |
| Zhen et al(b), 2020[27] | The USA | 108 | Panther Fusion | 58 | 0 | 0 | 50 |
| Lieberman et al, 2020[29] | The USA | 141 | Panther Fusion | 25 | 0 | 2 | 29 |
| Hogan et al, 2020[30] | The USA | 180 | Panther Fusion | 76 | 2 | 1 | 101 |
| Zhen et al(a), 2020[26] | The USA | 104 | Simplexa | 51 | 0 | 0 | 53 |
| Lieberman et al, 2020[29] | The USA | 141 | Simplexa | 11 | 0 | 0 | 8 |
| Bordi et al, 2020[31] | Italy | 278 | Simplexa | 99 | 8 | 0 | 171 |
| Lieberman et al, 2020[29] | The USA | 141 | Cobas® | 19 | 0 | 1 | 20 |
| Pujadas et al, 2020[32] | The USA | 963 | Cobas® | 639 | 39 | 1 | 284 |
| Ishige et al, 2020[33] | Japan | 24 | Cobas® | 4 | 0 | 0 | 20 |
| Poljak et al, 2020[34] | The USA | 716 | Cobas® | 123 | 3 | 3 | 587 |
| Zhen et al(b), 2020[27] | The USA | 108 | Xpert Xpress | 57 | 0 | 1 | 50 |
| Lieberman et al, 2020[29] | The USA | 141 | Xpert Xpress | 13 | 0 | 0 | 13 |
| Wolters et al, 2020[35] | Netherlands | 88 | Xpert Xpress | 58 | 0 | 0 | 30 |
| Broder et al, 2020[36] | The USA | 35 | Xpert Xpress | 34 | 0 | 0 | 1 |
| Loeffelholz et al, 2020[37] | The USA | 481 | Xpert Xpress | 219 | 11 | 1 | 250 |
| Baek et al, 2020[38] | Korea | 154 | RT-LAMP | 14 | 2 | 0 | 138 |
| Yan et al, 2020[39] | China | 130 | RT-LAMP | 58 | 0 | 0 | 72 |
| Huang et al, 2020[40] | China | 16 | RT-LAMP | 8 | 0 | 0 | 8 |
| Broughton et al, 2020[41] | The USA | 82 | RT-LAMP | 38 | 0 | 2 | 42 |
| Lu et al(a), 2020[42] | China | 56 | RT-LAMP | 34 | 2 | 2 | 18 |
| Lu et al(b), 2020[43] | China | 24 | RT-LAMP | 17 | 0 | 0 | 17 |

### Quality evaluation

All eligible articles were assessed through the Quadas-2 tool. The result suggested that all articles exhibited good quality (Supplementary Figure 2).

### Threshold effect

The Spearman correlation coefficient and p-value were utilized to evaluate the threshold effect via Meta-DiSc software 1.4. The Spearman correlation and p-value of ePlex were 0.500, *P*= 0.667; Panther Fusion were −0.200, *P*= 0.800; Simplexa were −0.500, *P*= 0.667; Cobas^®^ were 0.316, *P*= 0.684; Xpert Xpress were 0.300, *P*= 0.624; RT-LAMP were −0.348, *P*= 0.499. According to these results, no significant threshold exist in these studies on evaluating the different methods in detecting COVID-19.

### Diagnostic efficiency of different methods

#### ePlex

Three articles reported this method were included. The results showed the pooled sensitivity was 0.94, 95%CI (0.89-0.98) (*I*^*2*^=24.9%, *P*=0.2641) (Figure 1A); pooled specificity was 1.0, 95%CI (0.97-1.0) (*I*^*2*^=0%, P=1.0)(Figure 1B); pooled PLR was 90.91, 95%CI (17.25-479.2) (*I*^*2*^=0%, *P*=0.9740) (Figure 1C); pooled NLR was 0.07, 95%CI (0.03-0.13) (*I*^*2*^=0%, *P*=0.5502) (Figure 1D); pooled DOR was 1409.49, 95%CI (202.69-9801.44) (*I*^*2*^=0%, *P*=0.9297) (Figure 1E); AUC=0.9979, Q*=0.9840 (Figure 1F).

**Figure 1.**
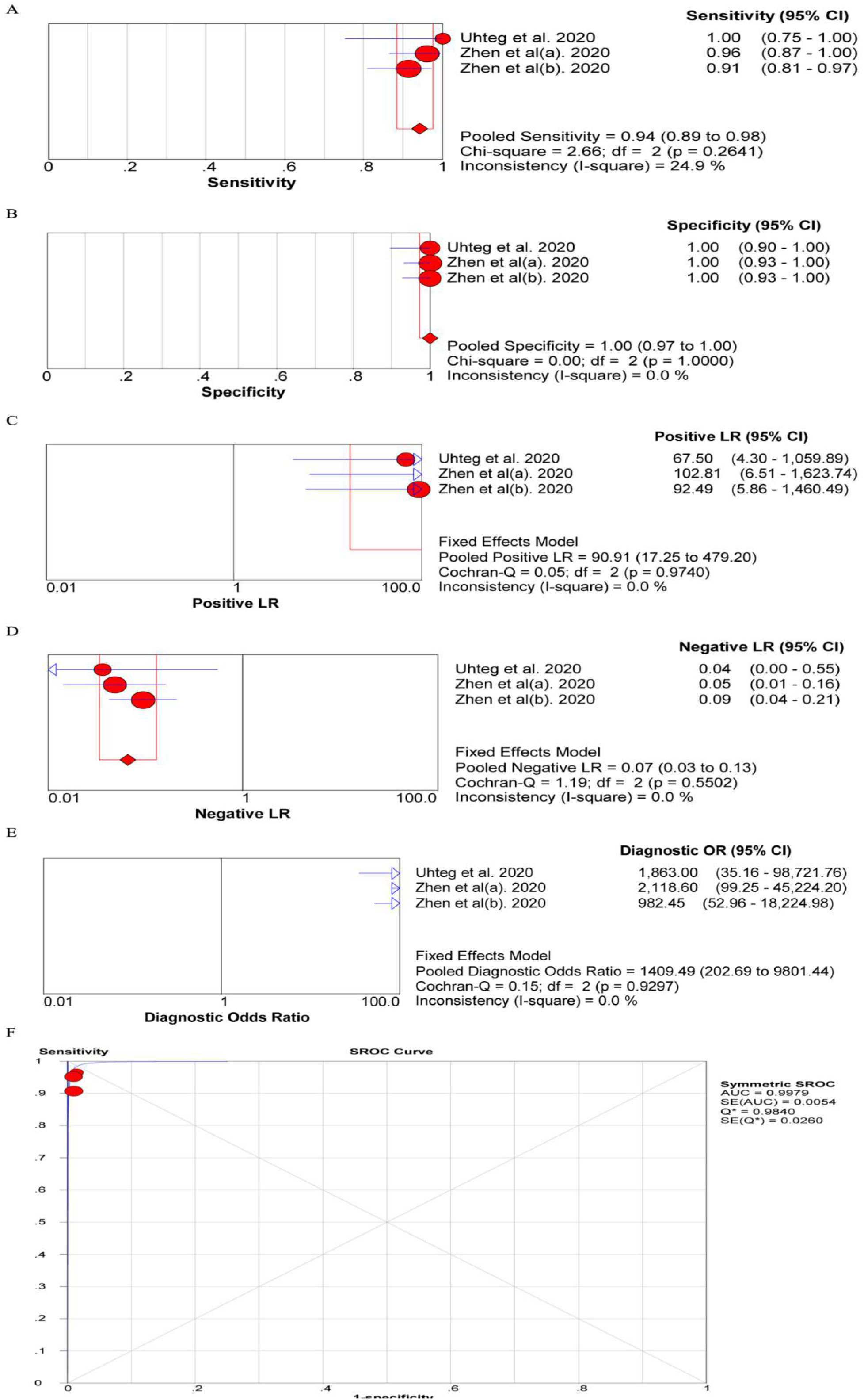
The diagnostic efficiency of ePlex: pooled summary sensitivity (A), specificity (B), PLR (C), NLR (D), DOR (E), sROC (F).

#### Panther Fusion

Four articles described this method were included. The results revealed that the pooled sensitivity was 0.99, 95%CI (0.96-1.0) (*I*^*2*^=54.5%, *P*=0.0858) (Figure 2A); pooled specificity was 0.98, 95%CI (0.96-1.0) (*I*^*2*^=20.2%, *P*=0.2888) (Figure 2B); pooled PLR was 42.46, 95%CI (18.50-97.43) (*I*^*2*^=0%, *P*=0.6347) (Figure 2C); pooled NLR was 0.02, 95%CI (0.01-0.11) (*I*^*2*^=54.9%, *P*=0.0839) (Figure 2D); pooled DOR was 2300.38, 95%CI (493.51-10722.63) (*I*^*2*^=0%, *P*=0.6673) (Figure 2E); AUC=0.9970, Q*=0.9799 (Figure 2F).

**Figure 2.**
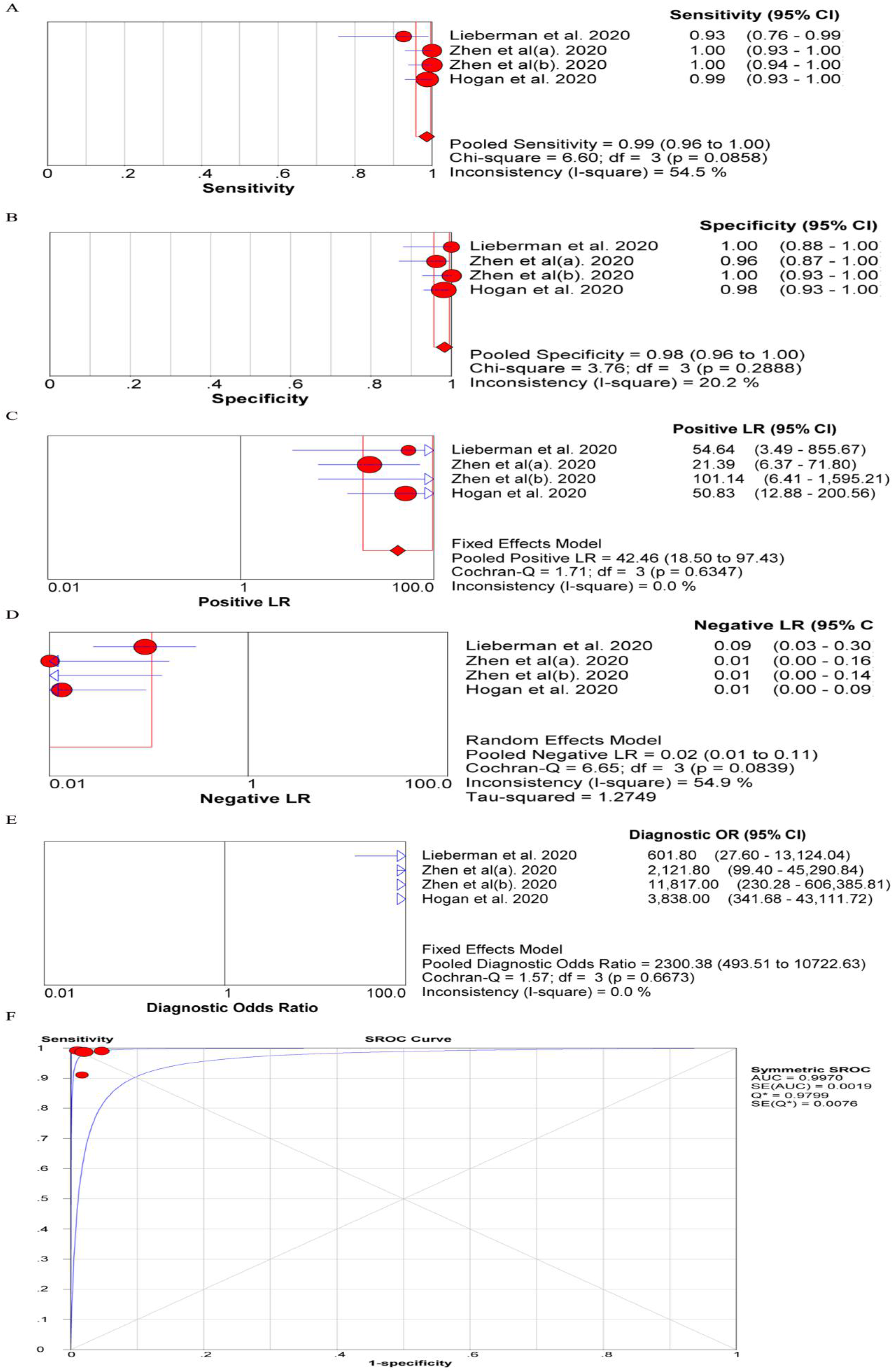
The diagnostic efficiency of Panther Fusion: pooled summary sensitivity (A), specificity (B), PLR (C), NLR (D), DOR (E), sROC (F).

#### Simplexa

Three articles exhibited this method were included. The results demonstrated that the pooled sensitivity was 1.0, 95%CI (0.98-1.0) (*I*^*2*^=0%, *P*=1.0) (Figure 3A); pooled specificity was 0.97, 95%CI (0.94-0.99) (*I*^*2*^=58.2%, *P*=0.0914) (Figure 3B); pooled PLR was 26.67, 95%CI (13.94-51.03) (*I*^*2*^=0%, *P*=0.4563) (Figure 3C); pooled NLR was 0.01, 95%CI (0.00-0.05) (*I*^*2*^=0%, *P*=0.4677) (Figure 3D); pooled DOR was 3100.93, 95%CI (406.14-23829.04) (*I*^*2*^=0%, *P*=0.4844) (Figure 3E); AUC=0.9970, Q*=0.9800 (Figure 3F).

**Figure 3.**
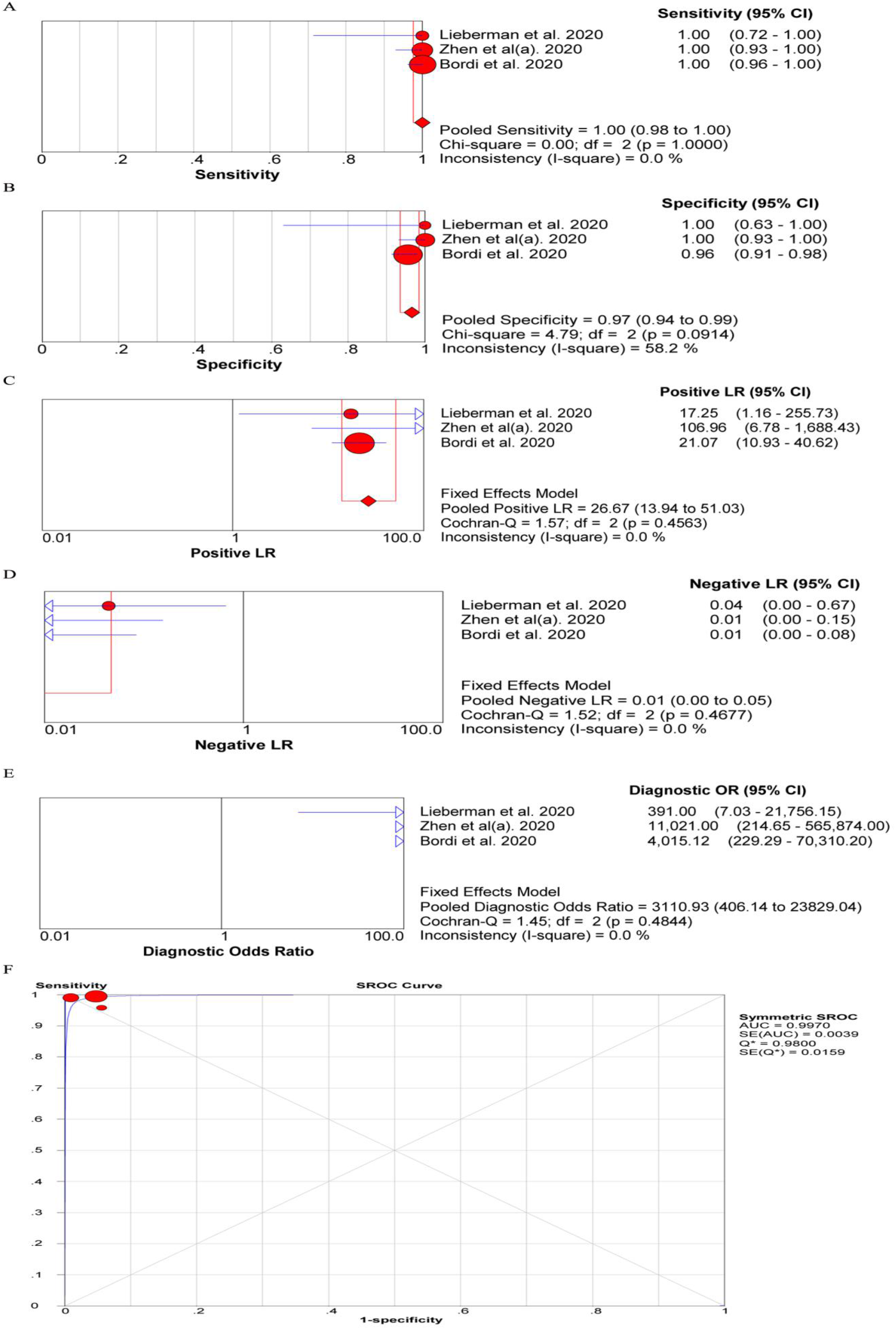
The diagnostic efficiency of Simplexa: pooled summary sensitivity (A), specificity (B), PLR (C), NLR (D), DOR (E), sROC (F).

#### Cobas^®^

Four articles described this method were included. The results demonstrated that the pooled sensitivity was 0.99, 95%CI (0.99-1.0) (*I*^*2*^=68%, *P*=0.0247) (Figure 4A); pooled specificity was 0.96, 95%CI (0.94-0.97) (*I*^*2*^=95.6%, *P*=0.000) (Figure 4B); pooled PLR was 37.82, 95%CI (4.6-311.22) (*I*^*2*^=90.3%, *P*=0.000) (Figure 4C); pooled NLR was 0.02, 95%CI (0.00-0.13) (*I*^*2*^=75.6%, *P*=0.0064) (Figure 4D); pooled DOR was 3754.05, 95%CI (1047.91-13448.55) (*I*^*2*^=15.1%, *P*=0.3163) (Figure 4E); AUC=0.9973, Q*=0.9810 (Figure 4F).

**Figure 4.**
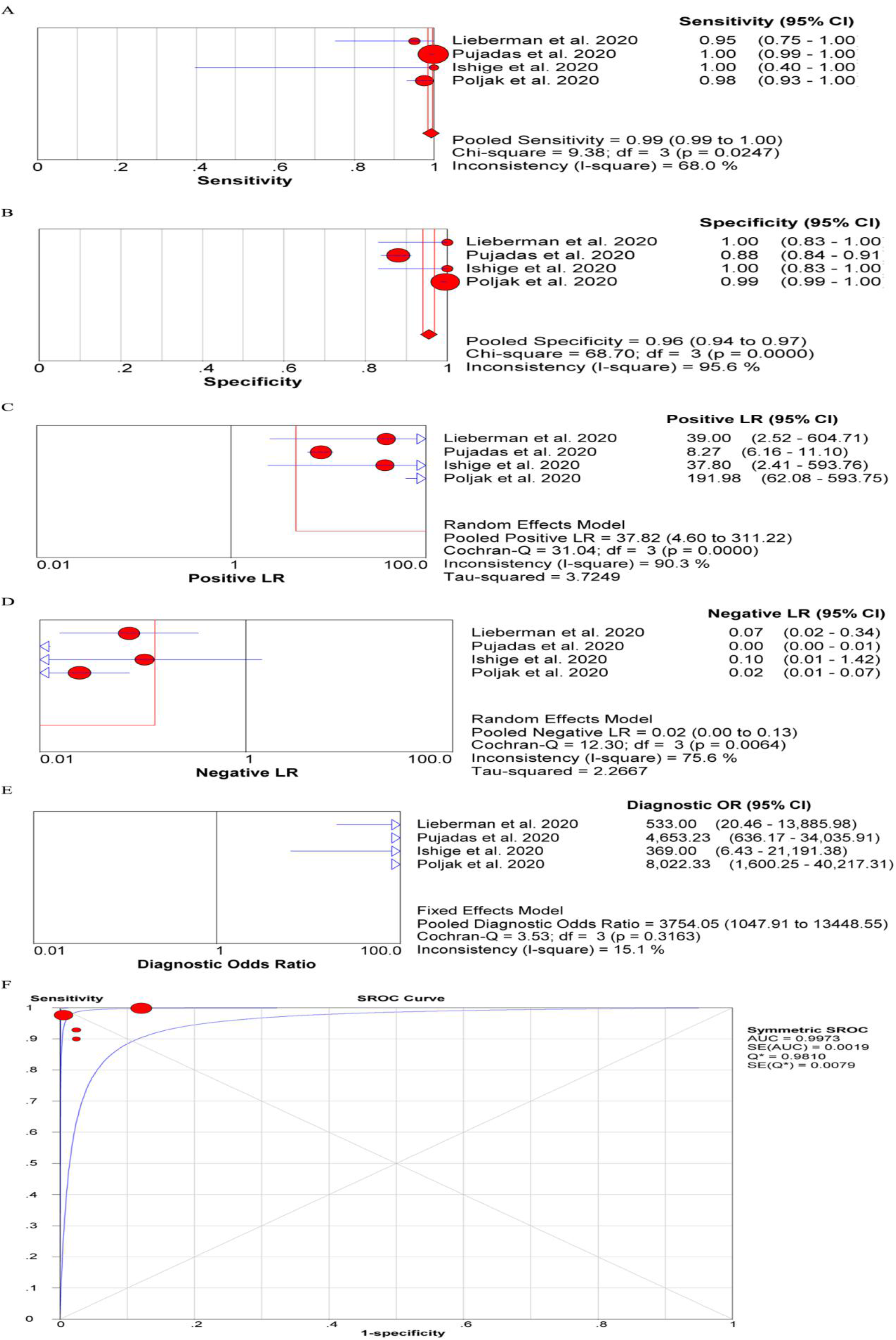
The diagnostic efficiency of Cobas^®^: pooled summary sensitivity (A), specificity (B), PLR (C), NLR (D), DOR (E), sROC (F).

#### Xpert Xpress

Five articles reported this method were included. The results revealed that the pooled sensitivity was 0.99, 95%CI (0.98-1.00) (*I*^*2*^=0%, *P*=0.7132) (Figure 5A); pooled specificity was 0.97, 95%CI (0.95-0.98) (*I*^*2*^=42%, *P*=0.1417) (Figure 5B); pooled PLR was 27.44, 95%CI (16.00-47.06) (*I*^*2*^=0%, *P*=0.4150) (Figure 5C); pooled NLR was 0.01, 95%CI (0.00-0.03) (*I*^*2*^=0%, *P*=0.5535) (Figure 5D); pooled DOR was 3488.15, 95%CI (868.18-14014.59) (*I*^*2*^=0%, *P*=0.6383) (Figure 5E); AUC=0.9977, Q*=0.9829 (Figure 5F).

**Figure 5.**
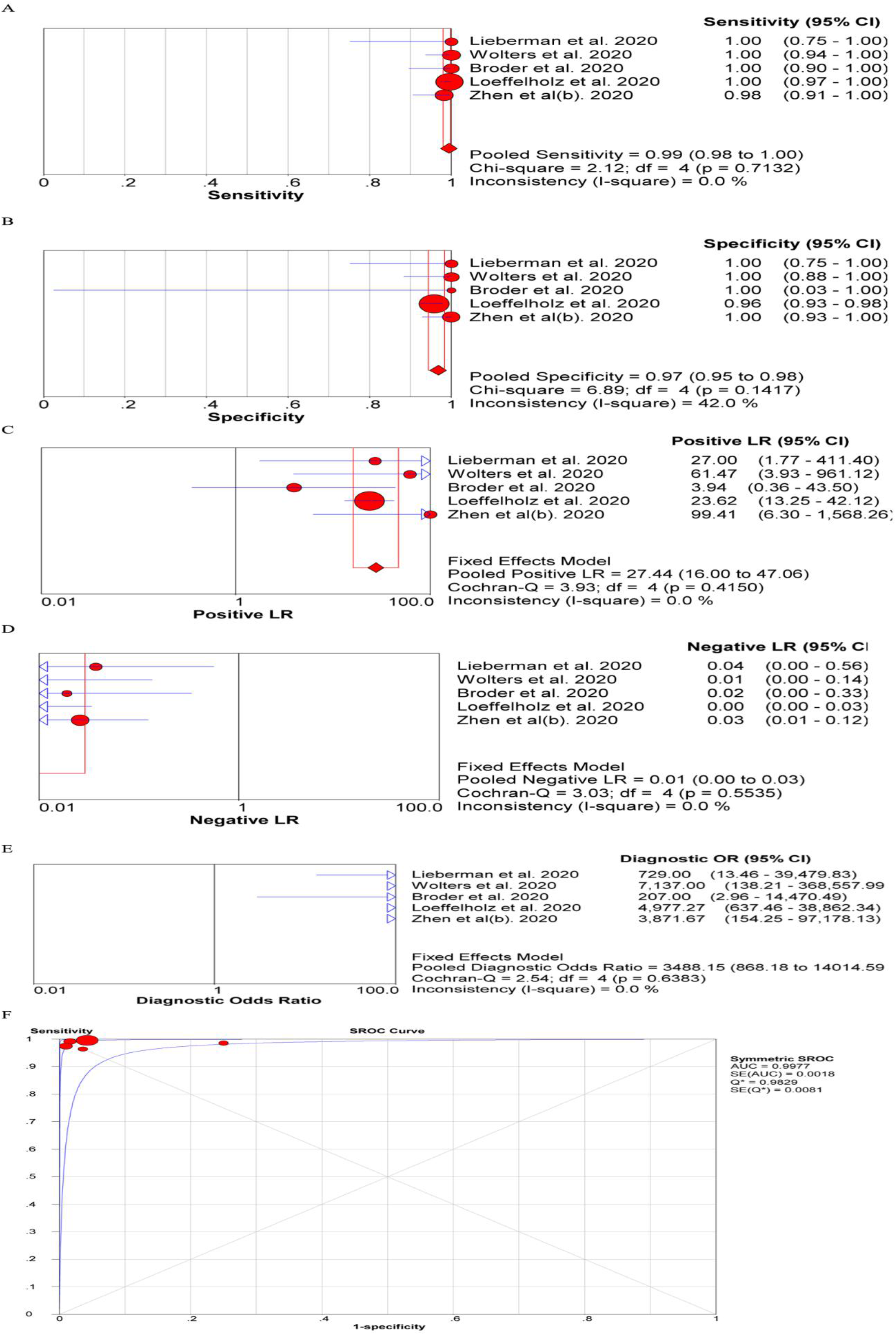
The diagnostic efficiency of Xpert Xpress: pooled summary sensitivity (A), specificity (B), PLR (C), NLR (D), DOR (E), sROC (F).

#### RT-LAMP

Six articles exhibited this method were included. The results suggested that the pooled sensitivity was 0.98, 95%CI (0.94-0.99) (*I*^*2*^=25.5%, *P*=0.2429) (Figure 6A); pooled specificity was 0.99, 95%CI (0.97-1.0) (*I*^*2*^=41.1%, *P*=0.1312) (Figure 6B); pooled PLR was 36.22, 95%CI (16.11-81.40) (*I*^*2*^=16.9%, *P*=0.3045) (Figure 6C); pooled NLR was 0.04, 95%CI (0.02-0.08) (*I*^*2*^=0%, *P*=0.7542) (Figure 6D); pooled DOR was 751.24, 95%CI (227.79-2477.54) (*I*^*2*^=7%, *P*=0.3716) (Figure 6E); AUC=0.9905, Q*=0.9596 (Figure 6F).

**Figure 6.**
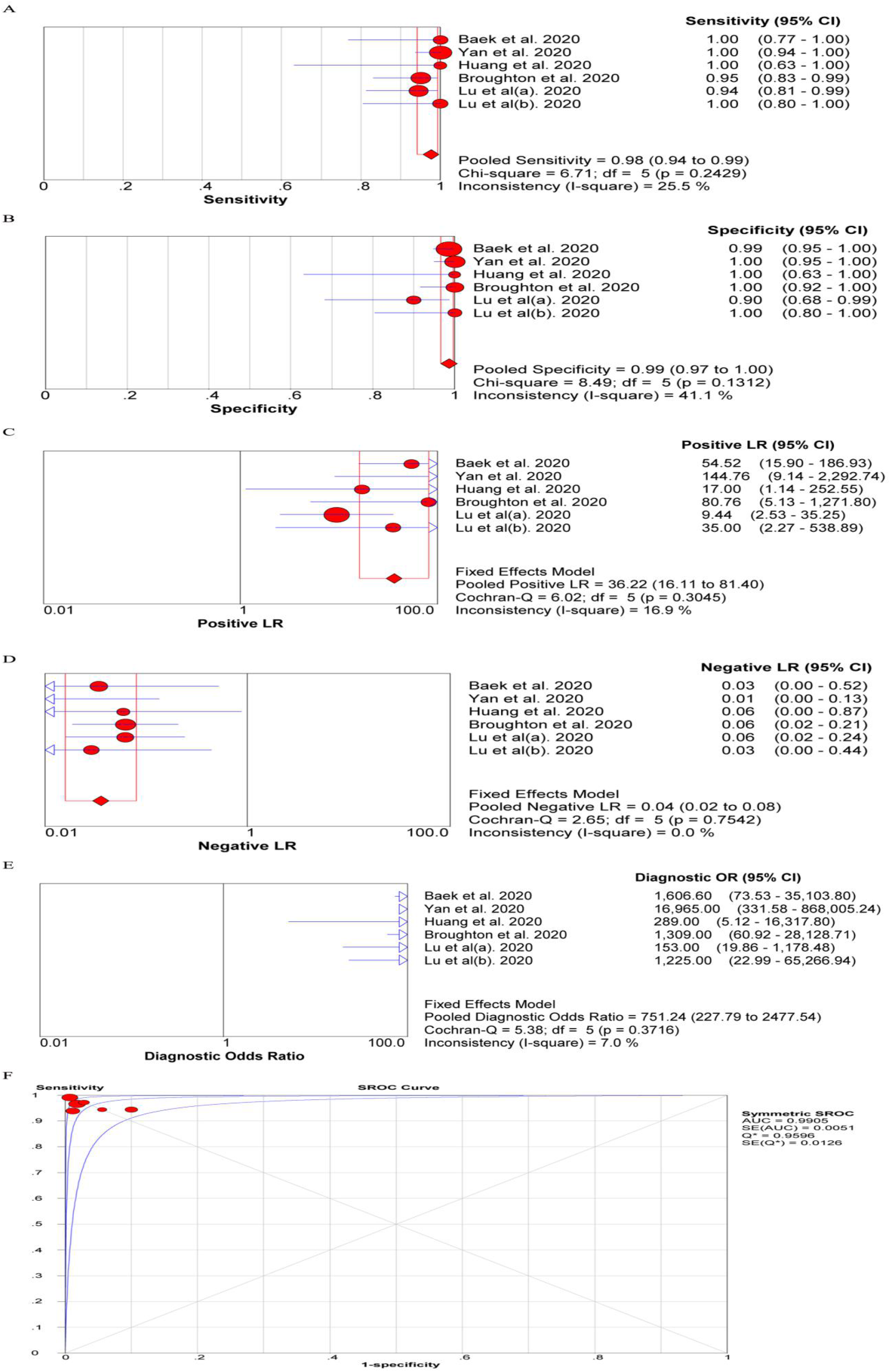
The diagnostic efficiency of RT-LAMP: pooled summary sensitivity (A), specificity (B), PLR (C), NLR (D), DOR (E), sROC (F).

#### Publication bias

The funnel plot exhibited that no significant publication bias in analysis on ePlex, Panther Fusion, Simplexa, RT-LAMP, Cobas^®^ and Xpert Xpress, because of the relatively symmetrical in this funnel plot (all P-value >0.1)(Supplementary Figure 3).

## Discussion

The COVID-19 pandemic is putting enormous pressure on clinical and public health laboratories. The COVID-19 pandemic may be caused by widespread transmission, viral detection is key to isolate positive patients and stop viral transmission(44-48). rRT-PCR is the most reliable and widely used technology to diagnose viruses including coronaviruses(44-48). However, rRT-PCR has some limitations, which can not meet the huge demand for the global pandemic of COVID-19. Recently, the US FDA has been authorized multiple rapid molecular tests to meet the huge diagnostic need. Different diagnostic methods have proposed different virus targets for detection of the virus, which would detect SARS-CoV-2 and other related beta coronaviruses such as SARS-CoV, including RNA-dependent RNA polymerase (RdRp), envelope (E), spike (S), Open Reading Frame (ORF) 1a and nucleocapsid (N)(21, 47, 51, 52). This article discusses some of the diagnostic methods include EUA-granted assays.

To date, this study was the first meta-analysis on systematically evaluating the diagnostic efficiency of different method for detecting COVID-19. In this study, we analyzed the pooled sensitivity, specificity, PLR, NLR, DOR, AUC and Q* on each methods (ePlex, Panther Fusion, Simplexa, Cobas^®^, Xpert Xpress and RT-LAMP), respectively. The results demonstrated that these above methods bear higher sensitivity and specificity, and might be efficient methods complement to the gold standard.

The ePlex assay targets the N gene of SARS-CoV-2, which is an in vitro diagnostic test. The ePlex has a relatively short turnaround time, simple operational flow, but a shortage of supply and inventory limits its full implementation.

The Panther Fusion SARS-CoV-2 assay targets two conserved regions of ORF1ab in the same fluorescence channel. This platform is automated, high-throughput systems that can process >1,000 specimens in 24 hours, which is met the huge diagnostic need.

Simplexa COVID-19 Direct assay targeted two distinct regions of the SARS-COV-2 genome, the surface (S) gene and the open reading frame 1AB(ORF1ab), distinguish with FAM and JOE fluorescent probes. Compared with RT-PCR, which requires nucleic acid extraction from clinical samples, Simplexa COVID-19 Direct assay can be detected directly from clinical samples. This detection method is easy to operate and does not require additional equipment such as a centrifuge or extraction system, which indicated is promising for laboratory diagnosis of COVID-19 and field application.

Cobas^®^ SARS-CoV-2 is a qualitative dual target assay, includes the ORF1/a nonstructural regional unique to SARS-CoV-2 and a region on the E gene, which is conserved across the sarbecovirus subgenus. Cobas^®^ SARS-CoV-2 is based on fully automated sample preparation (nucleic acid extraction and purification) followed by PCR amplification and detection(53). Automated solutions for molecular diagnostics can help process large numbers of samples, save testing time, allow non - professional operators. The assay has passed clinical evaluation and received EUA from the U.S. FDA(54-56).

The Xpert Xpress SARS-CoV-2 assay targets two genes, the E-gene (Sarbeco specific) and N2-gene (SARS-CoV-2 specific), which received EUA status on March 20, 2020. The Xpert test platform integrates specimen processing, nucleic acid extraction, reverse transcriptase polymerase chain reaction amplification of SARS-CoV-2 RNA, and amplicon detection in a single cartridge, which improves actionability overall. This assay is simple to operate with the least technical interventions, and FDA has authorized trained non-laboratorians to test.

RT-LAMP methodology is regarded as a new generation diagnostics(57), which was developed by Notomi et al.in 2000(58). This method has high sensitivity, high specificity, simple method, low cost and time saving, which has been widely applied for the detection of influenza virus, MERS-CoV, West Nile virus, Ebola virus, Zika virus, yellow fever virus, and a variety of other pathogens(59-64). The detection results are based on colorimetric display is easy to understand and does not require any expensive equipment which is suitable for countries with limited laboratory capacity.

We also encountered some limitations: i) the sample size of each method for detecting COVID-19 was relatively small. ii) the heterogeneity of some studies was significant though we have divided this meta-analysis based on the different methods, which might affect the stability of results to some extent. Therefore, more well designed study involving the different methods with a large sample size is urgent to be conducted.

Summarily, these methods (ePlex, Panther Fusion, Simplexa, Cobas^®^, Xpert Xpress and RT-LAMP) bear higher sensitivity and specificity, and might be efficient methods complement to the gold standard.

## Data Availability

Not applicable

## Acknowledgments

none

## Access to data

not applicable

## Contribution

Haitao Yang, Yuzhu Lan and Baosong Xie designed this study. The searching and data extracted were performed by Haitao Yang and Yuzhu Lan. Xiujuan Yao and sheng Lin analyzed the data, then created tables and figures. Haitao Yang and Yuzhu Chen drafted the manuscript. Xiujuan Yao and sheng Lin revised this manuscript. all authors commented on previous versions of the manuscript and approved the final manuscript.

---

**Supplementary Figure 1.**
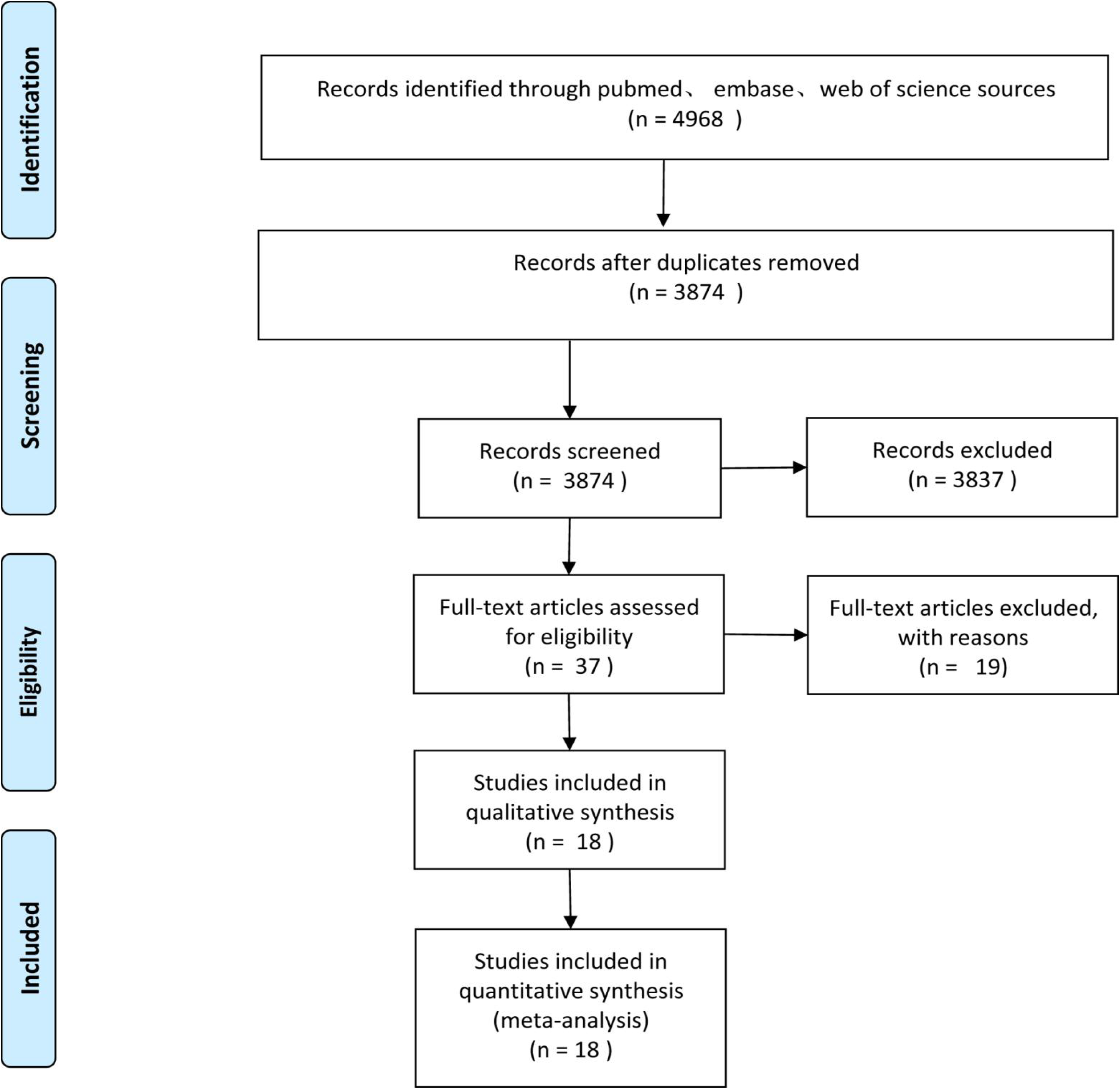
The flow chart for study selection.

**Supplementary Figure 2.**
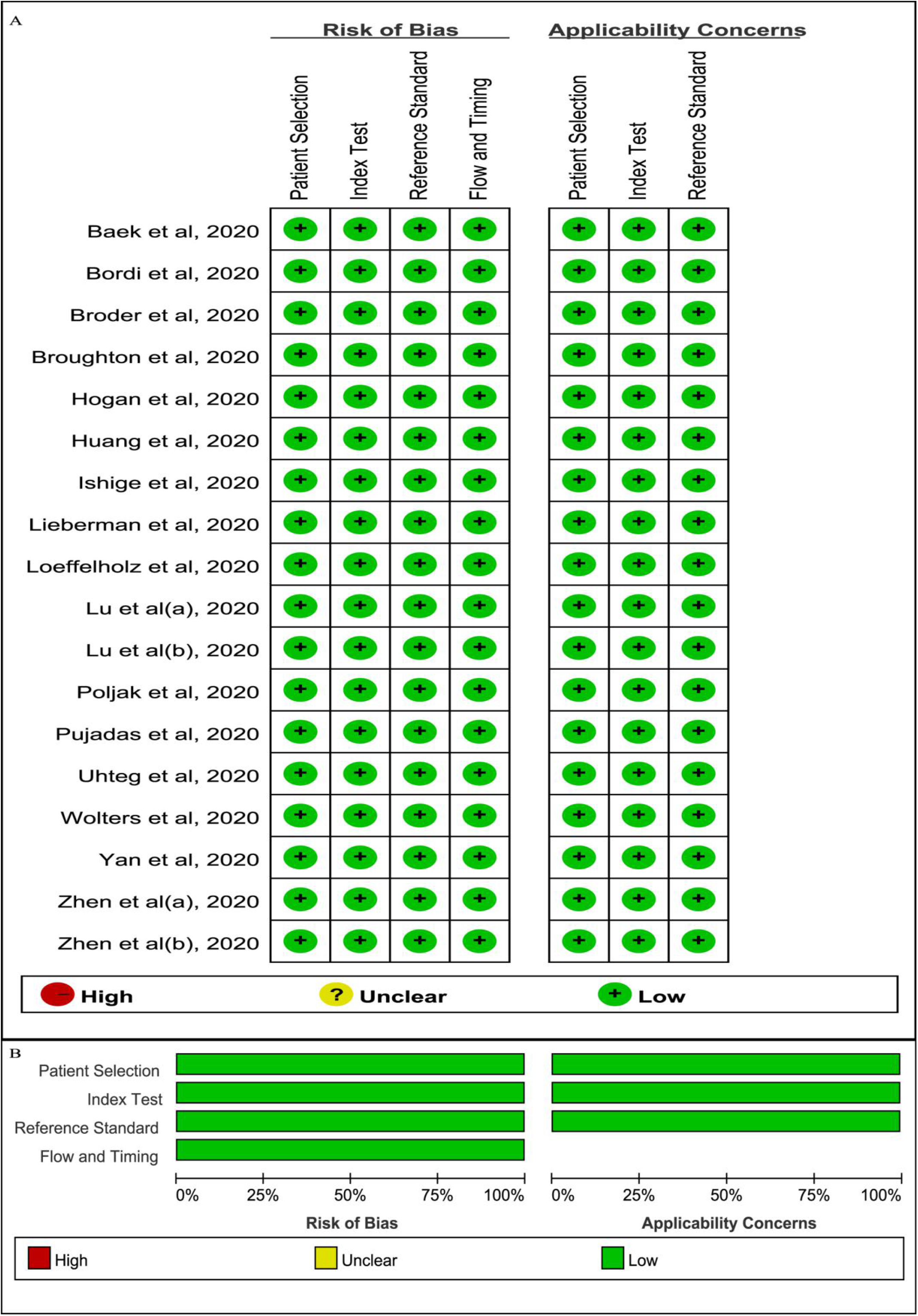
Quality assessment of included studies by Quadas-2: summary table of rating of risk of bias and applicability concerns for each study (A); cumulative barplot of risk of bias and applicability concerns across all studies (B).

**Supplementary Figure 3.**
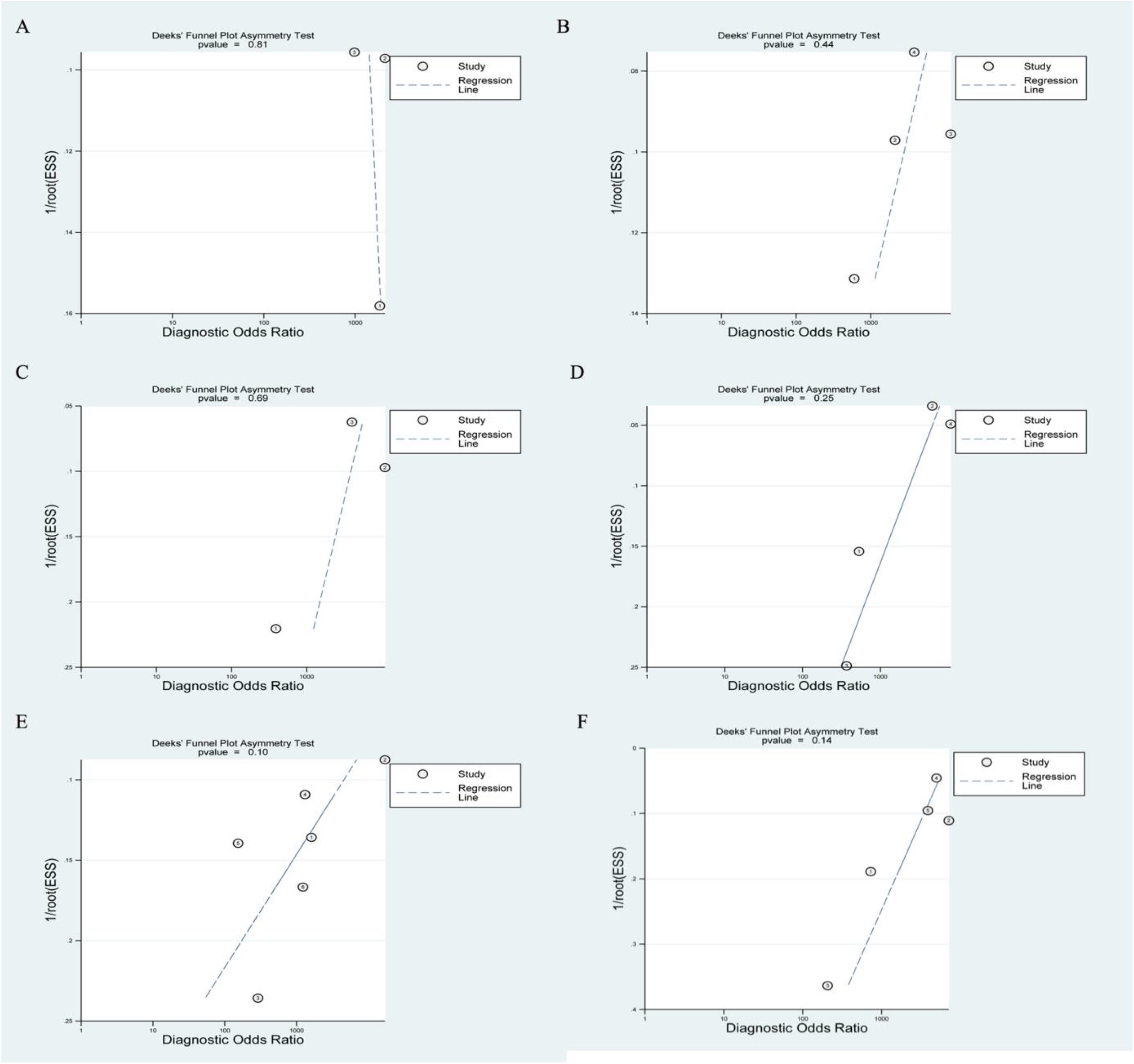
Plot of Deeks asymmetry test for publication bias in different methods: ePlex (A); Panther Fusion (B); Simplexa (C); Cobas^®^ (D); RT-LAMP (E); Xpert Xpress (F).

